# Blood Plasma Metabolomics to Support Uveal Melanoma Diagnosis

**DOI:** 10.1101/2022.09.14.22279822

**Authors:** Daniël P. de Bruyn, Michiel Bongaerts, Ramon Bonte, Jolanda Vaarwater, Magda A. Meester-Smoor, Robert M. Verdijk, Dion Paridaens, Nicole C. Naus, Annelies de Klein, George J.G. Ruijter, Emine Kiliç, Erwin Brosens, the Rotterdam Ocular Melanoma Study (ROMS) Group

**Author notes:** Corresponding author: E. Brosens, PhD., Address: Dr. Molewaterplein 40, 3015GD Rotterdam; P.O. Box 2040, 3000CA Rotterdam, The Netherlands. Shared first author (Daniël P. de Bruyn, Michiel Bongaerts). Shared last author (Emine Kiliç, Erwin Brosens).

## Abstract

**Importance:** Uveal Melanomas (UM) micro-metastasis can be present prior to diagnosis and relapse after treatment. Earlier detection resulted in an increased incidence of small (T1 and T2) tumors allowing for novel eye-preserving treatment strategies but reducing available tumor tissue needed for prognostic genomic profiling, creating the need for minimal-invasive detection and novel prognostication methods.

**Objective:** To determine whether tumor presence can be confirmed using metabolite patterns in blood plasma and to evaluate if these patterns differ between high risk (BRCA1-associated protein-1, *BAP1*), intermediate risk (Splicing Factor 3b Subunit 1, *SF3B1*) and low risk (Eukaryotic Translation Initiation Factor 1A X-Linked, *EIF1AX*) mutated tumors.

**Design:** Retrospective observational study including discovery (n=53) and replication (n=42) convenience sample sets compared to unaffected control-participants (n=46) as well as across mutation-based subgroups.

**Setting:** Patients from two tertiary referral centers specialized in ocular oncology: The Rotterdam Eye Hospital and the Erasmus MC Cancer Institute were included.

**Participants:** Sex-matched controls and patients were included based on their prognostic relevant secondary driver mutations. Peripheral blood plasma was collected at diagnosis, prior to treatment. Exclusion criteria were the presence of other malignancies or co-occurrence of systemic diseases at time of diagnosis.

**Main outcome and measure:** Metabolite profiles of patients and control-participants were generated as mass/charge (m/z) features using ultra-high performance liquid chromatography mass-spectrometry. After normalization, discriminatory feature patterns were determined using a random forest classifier and leave-one-out cross-validation.

**Results:** We detected differential metabolic patterns with a sensitivity of 0.95 and 0.90 and a specificity of 0.98 and 0.98 in the positive and negative ion modes, respectively. The accuracy of the model for classifying the subgroups was insufficient for the discovery (0.600 and 0.614 in the positive and negative ion modes, respectively) and replication cohort (0.544 and 0.672 in the positive and negative ion modes, respectively).

**Conclusion and relevance:** Minimally invasive metabolomics does not discriminate between the prognostic relevant *BAP1, SF3B1* and *EIF1AX* mutated UM-subgroups. However, this technique has the potential to allow for minimal invasive screening as it distinguishes metabolite patterns in peripheral blood derived plasma of UM-patients from control-participants.

**Key points:** *Question:* Can we discriminate uveal melanoma patients and mutation subgroups from unaffected control-participants using the metabolome of peripheral blood plasma taken at time of diagnosis?

*Findings:* In this retrospective observational study, we find a low sensitivity and specificity to detect subgroups but a high sensitivity and specificity to discriminate patients from control-participants by measuring metabolite abundancy in plasma using ultra-high performance liquid chromatography mass-spectrometry and reach a receiver operating characteristic area under the curve of 0.993.

*Meaning:* These results suggest that surveying the metabolome of uveal melanoma patients could aid in the minimal invasive detection of uveal melanoma.

## Introduction

Uveal melanoma (UM) affects 2-9 per million persons yearly and improved detection methods lead up to earlier diagnoses and increased detection of small lesions^1,2^. Three mutually-exclusive secondary driver mutations in BRCA-associated protein 1 (*BAP1*), splicing factor 3 subunit B1 (*SF3B1*) and Eukaryotic translation initiation factor 1A, X-linked (*EIF1AX*) present with corresponding copy number variation profiles and are highly correlated to prognosis^3^ (eTable 1). Treatment of primary lesions has shifted from surgical intervention towards eye-preserving therapies^1,4^. Whilst preserving the patients’ eye, this strategy generally reduces available tumor material for molecular prognostication. Therefore, other prognostic modalities need to be explored. Blood-based biomarkers have the potential to replace invasive characterization, but tumor biomarker abundance in blood of UM-patients is often low^4^. Untargeted metabolomics allows for hypothesis-free biomarker detection and plasma metabolome studies show promising results in the detection of different types of cancer ^5,6^. We investigated whether differences in metabolite patterns could distinguish UM-patients from control-participants and if we could identify prognostically relevant UM-patient subgroups.

## Methods

In this retrospective observational study, we determined metabolite abundancies obtained from peripheral blood. This study adhered to the tenets of the Declaration of Helsinki. Ethical approval was obtained from the local Medical Ethics Committee (MEC) of the Erasmus MC Rotterdam. We retrospectively included samples of UM-patients diagnosed between 1998 and 2021 (MEC-2009-375). Control-participants received care for cataract between 2016 and 2017 and samples were taken from the Combined Ophthalmic Research Rotterdam biobank (CORRBI) (MEC-2012-031, eTable 2 and 3). UM-patients and control-participants were excluded if malignancies or systemic diseases co-occurred. Minimally-invasive metabolomics was performed in a discovery and replication cohort in which technical and biological replicates were included to assess batch-effects (eFigure 1).

Statistical analyses were conducted with Python, v3.6.4 (Scipy, v1.5.2, and Scikit-learn, v.0.23.2) and GraphPad Prism 9, v9.2.0. Additional information about the sample collection and storage, metabolomics, statistical analyses, mass/charge features and annotations can be found in eMethods.

## Results

Clinical and demographic data of 156 participants in the discovery (n=114) and replication (n=42) cohorts are summarized in Table 1. UM-patients and control-participants were comparable regarding sex-distribution, comorbidities, and medication use. UM-subgroups were homogeneous regarding tumor size, location and primary driver mutation. Other prognostic factors were more prevalent in *BAP1*-mutated tumors (Table 1), conform the characteristics of these three UM-subclasses^3^. Multiple quality control approaches were used; biological and technical replicate analysis indicated good concordance and no systemic differences or batch effects were observed after normalization (eFigure 2, 3, 4, 5, 6 and eTable 3).

**Table 1.**
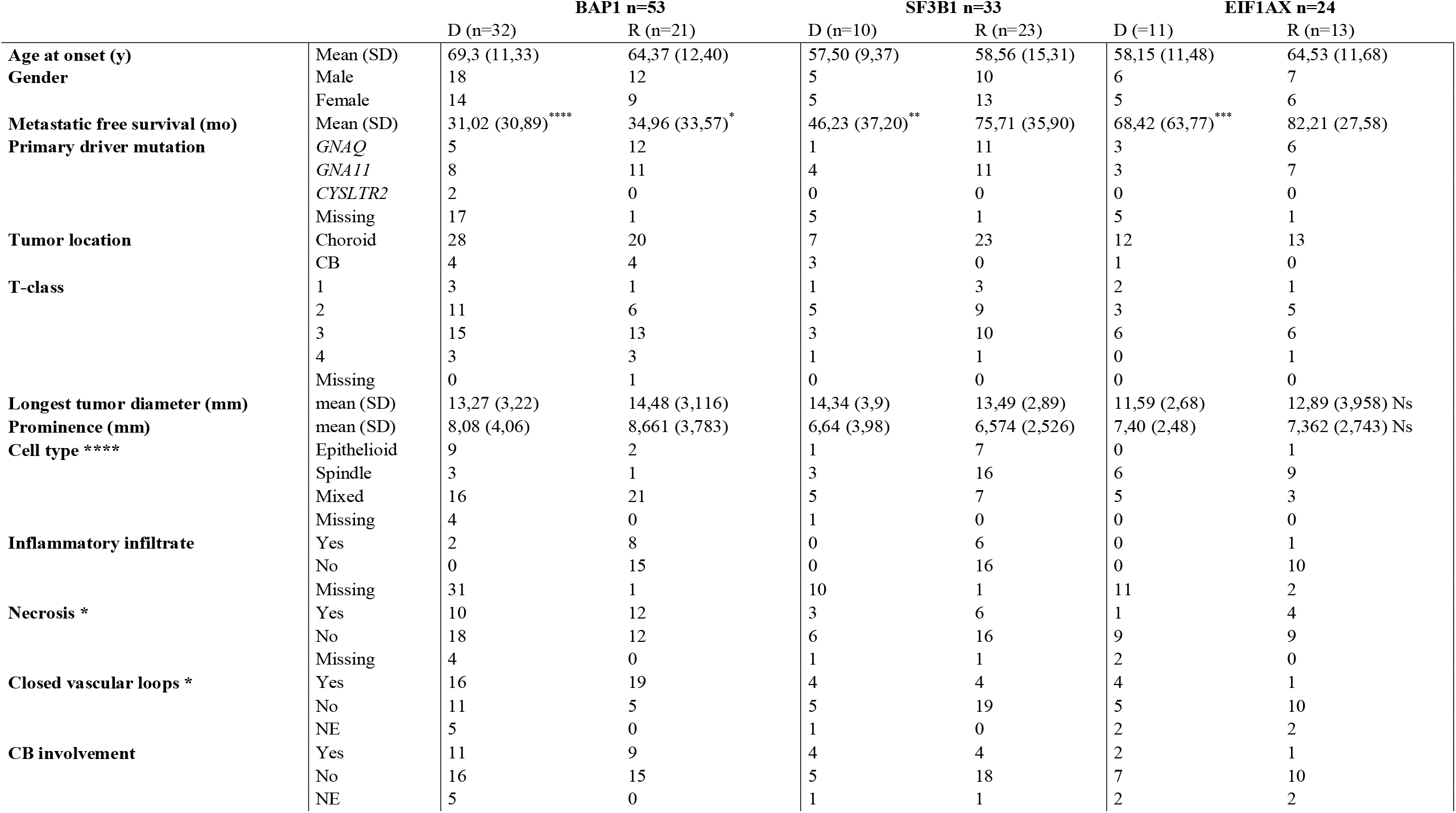
Cohort description. Cohort characteristics of discovery and replication cohorts used in this study. Significant differences between discovery and replication cohorts are depicted as followed: * p <0.05; ** p <0.005, *** p <0,001, **** p <0,0001. Patients are sex-matched with control-participants. The control-participants (n=46) have a mean age of 73.89 years (SD: 9.17) and include 21 males and 25 females. The control-participants were significantly older (p<0.0001). UM-subgroups are homogeneous in primary-driver mutation, tumor location and tumor size. Metastatic free survival is in months, longest tumor diameter and prominence are measured in millimeters. Abbreviations used: SD: standard deviation; CI: confidence interval; NE: not evaluated; Ns: non-significant difference, D: Discovery set, R: replication set; y: years; mo: months; mm: millimeters; CB: Ciliary body.

UM-patients have distinctly different metabolite patterns compared to control-participants. Metabolite profiles are analyzed in both ion modes to increase sensitivity, as metabolites can have an ionization preference for either protonation or deprotonation. A Random Forest classifier (RFC) was trained using a leave-one-out approach, classifying UM-patients versus control-participants. Our results show a sensitivity of 0.94 and 0.80 and specificity of 0.94 and 1.00 in positive and negative ion mode respectively for the discovery cohort (Table 2). Furthermore, for this analysis we obtained an area under the receiver operating characteristic curve of 0.993 (Figure 1).

**Table 2.**
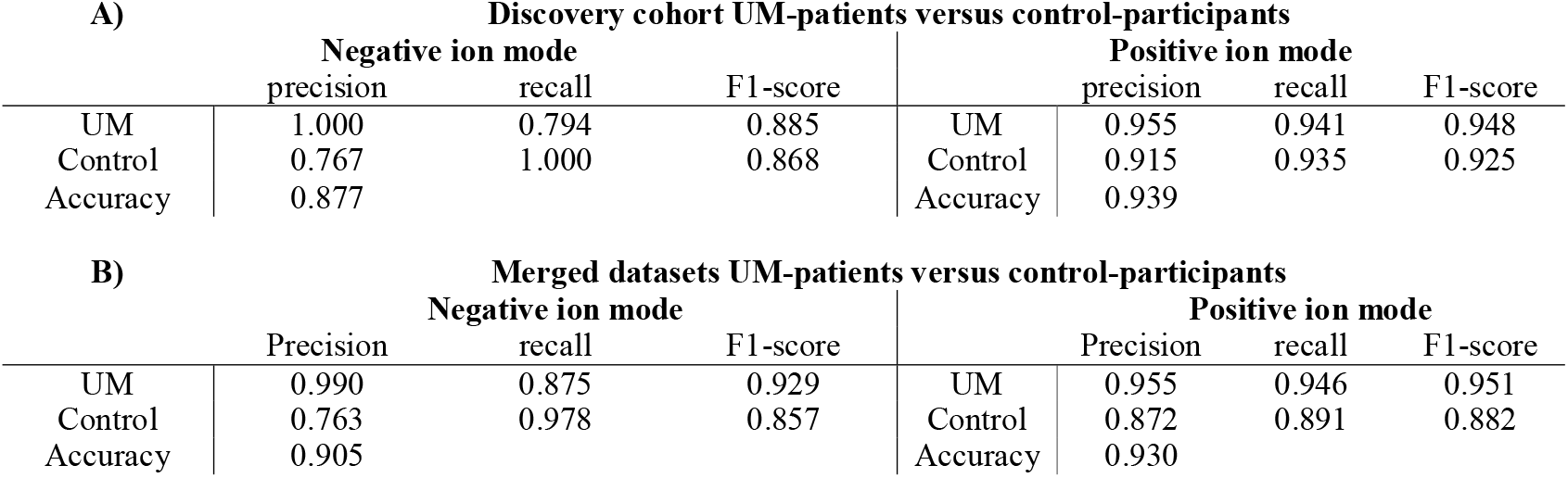
Validation of classifiers. Leave-one-out cross-validation of trained random forest classifiers that classify UM-samples versus control samples in the A) discovery cohort and B) merged dataset, as these datasets contained control samples. Accuracies of 0.88 and 0.94 are observed in the discovery cohort in the negative and positive ion modes, respectively. In the merged dataset the accuracies were 0.91 in the negative and 0.93 in the positive ion mode. Cross validation showed that the sensitivity to detect UM is 0.94 and 0.80 and the specificity is 0.94 and 1.00 in positive and negative ion modes in the discovery cohort and have become more robust after addition of validation samples.

| A) Discovery cohort UM-patients versus control-participants |  |  |  |  |  |  |  |
| --- | --- | --- | --- | --- | --- | --- | --- |
|  | Negative ion mode |  |  |  | Positive ion mode |  |  |
|  | precision | recall | F1-score |  | precision | recall | F1-score |
| UM | 1.000 | 0.794 | 0.885 | UM | 0.955 | 0.941 | 0.948 |
| Control | 0.767 | 1.000 | 0.868 | Control | 0.915 | 0.935 | 0.925 |
| Accuracy | 0.877 |  |  | Accuracy | 0.939 |  |  |

**Table 2. Validation of classifiers**
| B) Merged datasets UM-patients versus control-participants |  |  |  |  |  |  |  |
| --- | --- | --- | --- | --- | --- | --- | --- |
|  | Negative ion mode |  |  |  | Positive ion mode |  |  |
|  | Precision | recall | F1-score |  | Precision | recall | F1-score |
| UM | 0.990 | 0.875 | 0.929 | UM | 0.955 | 0.946 | 0.951 |
| Control | 0.763 | 0.978 | 0.857 | Control | 0.872 | 0.891 | 0.882 |
| Accuracy | 0.905 |  |  | Accuracy | 0.930 |  |  |

**Figure 1.**
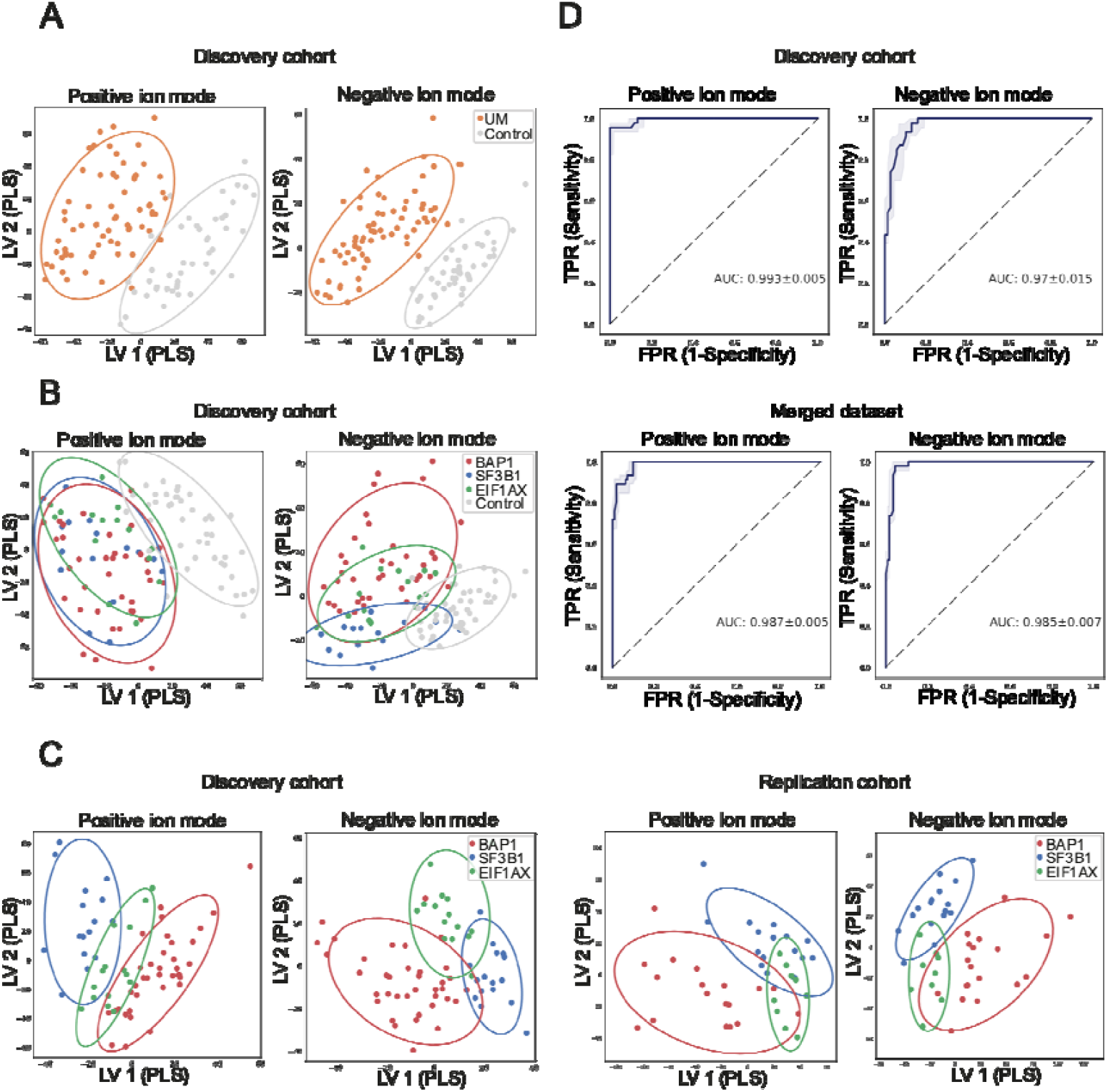
Distinguishing potential of metabolic patterns between UM-subtypes and near perfect differentiation between UM-patients and control-participants. Mass/charge features and retention times were determined using ultra-high performance liquid chromatography mass-spectrometry and these features were used for supervised dimensionality reduction methods and to train a Random Forest classifier (RFC). The partial least squares determinant analysis shows separation of UM-samples compared to samples of control-participants in the discovery cohort in the positive and negative ion modes (A). The control samples are separated when the three UM-subtypes are specified (B); and, individual UM-subtypes can be separated by metabolite profiles to some extent in the discovery and replication cohorts, respectively (C). However, training a random forest classifier to distinguish the different subgroups using a leave-one-out procedure led to poor classification performances (see eMethods). The receiver operating chart (ROC)-curve represents the performance of our RFC model by sensitivity on the Y-axis and 1-specificity on the X-axis for the discovery cohort and merged dataset, respectively (D). The overall test performance is shown by area under the curve (AUC) and an AUC greater than 0.7 is generally considered as a threshold for fair performance whereas an AUC of 1 indicates a perfect performance. The AUC of our test is 0.993 in the positive ion mode in the discovery cohort, which means a near perfect ability to differentiate metabolite patterns of UM-patients compared to control-participants. The AUC of 0.987 in the merged dataset indicates a robust differentiating power of our test when samples are added.

UMs can metastasize prior to diagnosis and consequently are still present after the primary tumor is removed or irradiated^3,7^. Micro-metastases can relapse and most patients harboring a *BAP1*-mutation will develop metastasizes within 60 months. Patients harboring an *SF3B1*-mutation will develop metastasis within 120 months^3^. We performed supervised dimensionality analyses (Figure 1) and trained an RFC to discriminate UM patients and the prognostic subgroups *BAP1, SF3B1* and *EIF1AX*. Unfortunately, the classification performance in the discovery and replication cohorts was insufficient to discriminate between subgroups (eTable 4). Similarly, we could not distinguish genetic subtypes associated with poor prognosis or formation of metastases (eTable 5). Feature abundancies are different in UM-patients compared to control-participants correlated with tumor size, however none are significantly correlated after multiple testing correction (eFigure 7). In-house annotated metabolites were used for pathway analysis and indicate upregulation of transfer (tRNA) charging and glycine degradation, and a downregulation of purine ribonucleosides degradation and the superpathway of citrulline metabolism (eFigure 8).

## Discussion

One of the hallmarks of cancer is metabolic reprogramming^8^. Differences in metabolite and protein abundancies in peripheral blood are used as biomarkers and can reveal patients harboring a malignancy^5,6,9,10^, detect upregulation of alternative energy consumption or inflammatory responses associated with cancer^5,9,10^. Here, we report a difference in plasma metabolite patterns between UM-patients and control-participants (Figure 1) and annotated, metabolites in UM-patients putatively associated with malignant processes (eFigure 8). This includes upregulation of tRNA-charging which promotes proliferation, metastasis, and invasion of malignant cells^11^. A lower abundance of arginine and citrulline is observed in line with upregulation of Arginase1, that catalyzes arginine hydrolysis, in plasma of UM-patients^9^. Furthermore, consistent with oncogenic processes, decreased adenine and guanine suggest either decreased purine degradation or increased salvage of purine bases. Several metabolite studies in UM focused on metastatic cell lines^12-14^. *BAP1*-mutated cell lines show upregulation of oxidative phosphorylation, pyruvate dehydrogenase and succinate dehydrogenase subunit A (SDHA)^12,13^. In lung cancer, the TCA-cycle is restored by upregulation of SDHA and valine metabolism^15^. Dysregulation of SDHA, oxidative phosphorylation and pyruvate dehydrogenase was not observed in patients’ plasma, but valine was less abundant in UM-patients compared to control-participants, consistent with an upregulation of the valine metabolism.

## Limitations

Unfortunately, not all metabolites are currently annotated with corresponding proteins, hampering translation from metabolites to proteins and biological pathways. Compared to other malignancies, UM are small and confined within the eye. Consequently, low biomarker levels in blood could be insufficient to discriminate subtypes. We evaluated after at least 60 months of clinical follow-up, but samples were taken at diagnosis. Perhaps the metabolic subgroup changes only present after a specific period and we measured before the effect of subtypes were detectable. We made use of an existing control cohort comprised of patients with ophthalmic diseases common in elderly. Additionally, control-participants were, on average, older than UM-patients (73.9 years vs. 63.0 years, p<0.0001). Blood samples of control-participants were stored shorter than five years, whereas storage times of samples of UM-patients ranged between 0 and 20 years. Although we normalized for technical variation, some of the difference between these cohorts could be due to technical differences and in future experiments the use of more similar, unaffected controls could strengthen the results.

## Conclusion

In UM, liquid biopsies can detect metastasis after measuring circulating tumor DNA and circulating tumor cells. With adequate levels, secondary driver mutations or copy number variation profiles can be derived. However, primary UMs are small and the amount of tumor-derived material in blood is low at diagnosis^4^. Metabolite patterns distinguish between UM-patients and control-participants at time of diagnosis and could thus be a valuable addition to other liquid biopsy approaches.

Additionally, serial phlebotomies are well tolerated and pose no risk, as opposed to intraocular biopsies. Previous studies report elevated metabolite levels involved in several oncogenic processes in plasma of cancer patients at the time of diagnosis. Here, pathway analysis of annotated metabolites showed several affected processes associated with malignancy, corresponding to prior studies. Potentially, untargeted metabolomics can detect multiple cancer-types and could be a valuable minimally-invasive screening method. Whilst future experiments could show further clinical applicability by comparing patients harboring small melanocytic intraocular lesions to distinguish benign nevi and nevi with malignant risk, this current study highlights the potential of untargeted metabolomics to support UM diagnosis.

## Data Availability

All data produced in the present study are available upon reasonable request to the authors

## Article information

### Corresponding Author

Erwin Brosens, PhD, Erasmus MC, PO box 2040, Ee-2089, 3000CA Rotterdam

### Author Contributions

Erwin Brosens and Michiel Bongaerts had full access to all the data in the study and takes responsibility for the integrity of the data and the accuracy of the data analysis.

Concept and design: Annelies de Klein, Emine Kiliç, Erwin Brosens. Acquisition, analysis, or interpretation of data: Ramon Bonte, Michiel Bongaerts, Jolanda Vaarwater, Daniel de Bruyn.

Drafting of the manuscript: Daniel de Bruyn, Michiel Bongaerts, Erwin Brosens. Critical revision of the manuscript for important intellectual content: All authors. Statistical analyses: Michiel Bongaerts, Daniel de Bruyn. Obtained funding: Daniel de Bruyn, Emine Kilic and Erwin Brosens.

Administrative, technical, or material support: Daniel de Bruyn, Ramon Bonte, Jolanda Vaarwater, Dion Paridaens, Nicole Naus, Emine Kilic. Supervision: Emine Kilic and Erwin Brosens

### Group information

The Rotterdam Ocular Melanoma Study Group (ROMS) is a collaborative research group with members from the Rotterdam Eye Hospital, Departments of Ophthalmology, Pathology and Clinical Genetics, of the Erasmus MC, Rotterdam, The Netherlands.

### Additional Contributions

We are grateful to all participants who took part in this study acknowledge the role of the clinicians, nurses, clinical coordinators, and investigators who collected material and clinical data on UM cases and control-participants.

