## Supplementary files for "Blood Plasma Metabolomics to Support Uveal Melanoma Diagnosis"

**Electronic media belonging to the manuscript:**

*^1^Department of Ophthalmology, ^2^Department of Clinical Genetics, ^3^Erasmus MC Cancer Institute, 3000 CA Rotterdam, The Netherlands, ^4^The Rotterdam Eye Hospital, 3011 BH Rotterdam, The Netherlands, ^5^Department of Pathology, Section Ophthalmic Pathology, Erasmus MC Rotterdam, 3000 CA Rotterdam, The Netherlands, ^6^Department of Pathology, Leiden University Medical Center, 2333 ZA Leiden, The Netherlands*

**Electronic media Content**

**eFigure**

1. Sample numbers in discovery, replication and merged datasets.
2. Quality control characteristics by internal standards in discovery set.
3. Quality control characteristics by internals standards in replication set.
4. Quality control by two-dimensionality reduction analyses.
5. Assessment of Metchalizer normalization in the merged datasets.
6. Assessment of technical and biological variation by using replicates in the discovery cohort and merged dataset.
7. Correlation of m/z features with tumor size and differential abundance between UM-patients and control-participants.
8. Pathway analysis of top-10 affected pathways in UM-patients compared to control-participants.

**eTable**

1. Association of copy number variations and secondary driver mutations with prognosis.
2. Patient characteristics of control-participants.
3. Patient and sample characteristics of technical and biological replicates.
4. No differentially abundant metabolite patterns in plasma of UM-patients between subtypes.
5. No differences in metabolite patterns associated with poor prognosis.

**eMethods**

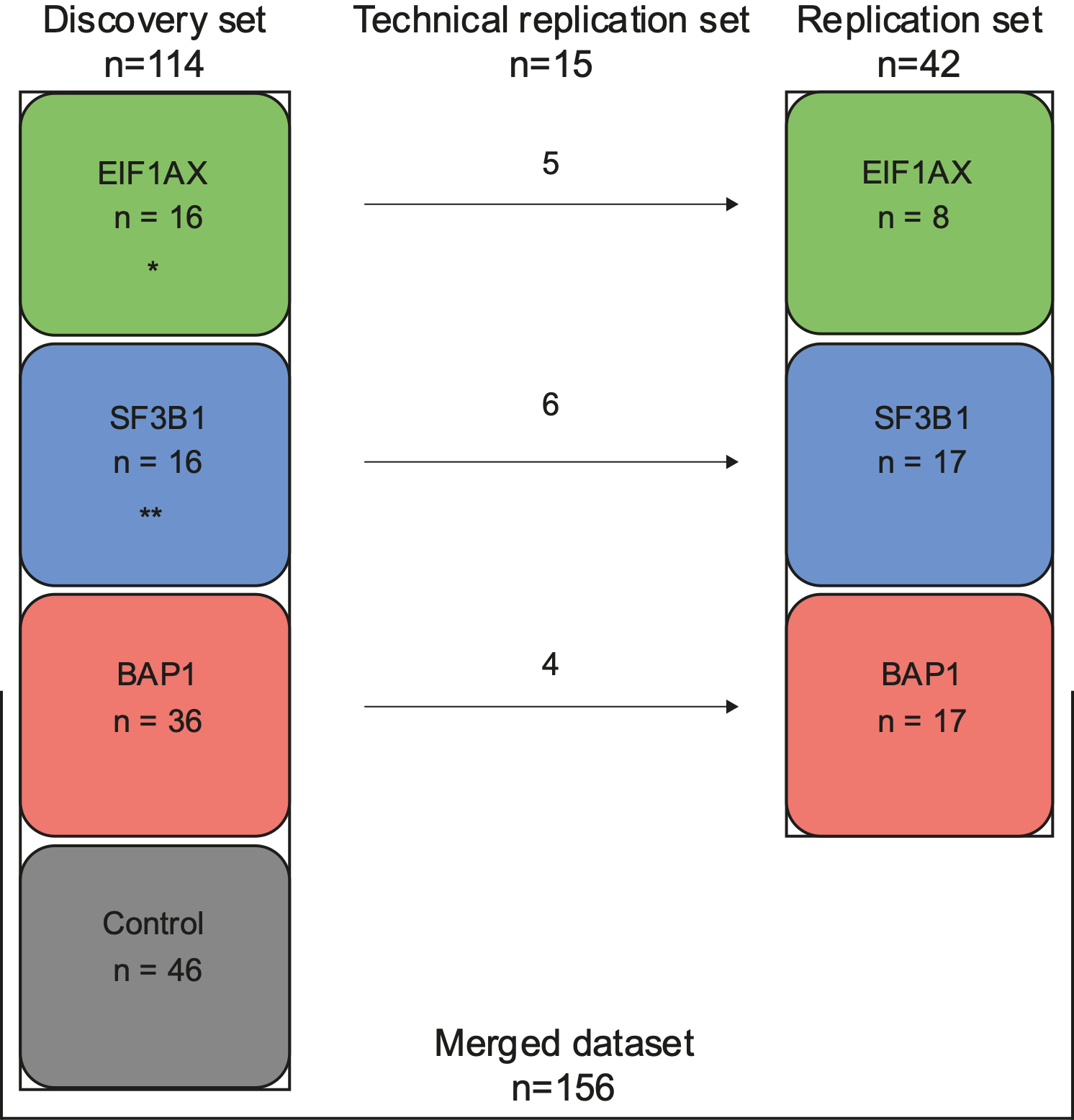

**eFigure 1.** **Sample numbers in discovery, replication and merged datasets.**

156 participants enrolled in our study. Our discovery set comprised of 16 patients harboring an *EIF1AX-*mutated tumor, 16 patients harboring an *SF3B1*-mutated tumor and 36 patients harboring a *BAP1*-mutated tumor. Additionally, 46 control-participants were included in the discovery set. The replication set included 8 patients harboring an *EIF1AX*-mutated tumor, 17 patients harboring an *SF3B1*-mutated tumor and 17 patients harboring a *BAP1*-mutated tumor. To assess batch effects in the merged dataset (discovery and replication set combined), 15 samples were used as a technical replication set (see eFigure 4). These technical replicates were used in the discovery cohort for exploring differential features (and subsequent metabolites). Afterwards, in the replication cohort, these samples were only used for quality control evaluation. Two patients harboring an *SF3B1-*mutated tumor and one patient harboring an *EIF1AX-*mutated tumor had an additional plasma sample retrieved at the same time-point included in the replication set as biological replicates to assess similarity in the metabolite profiles.

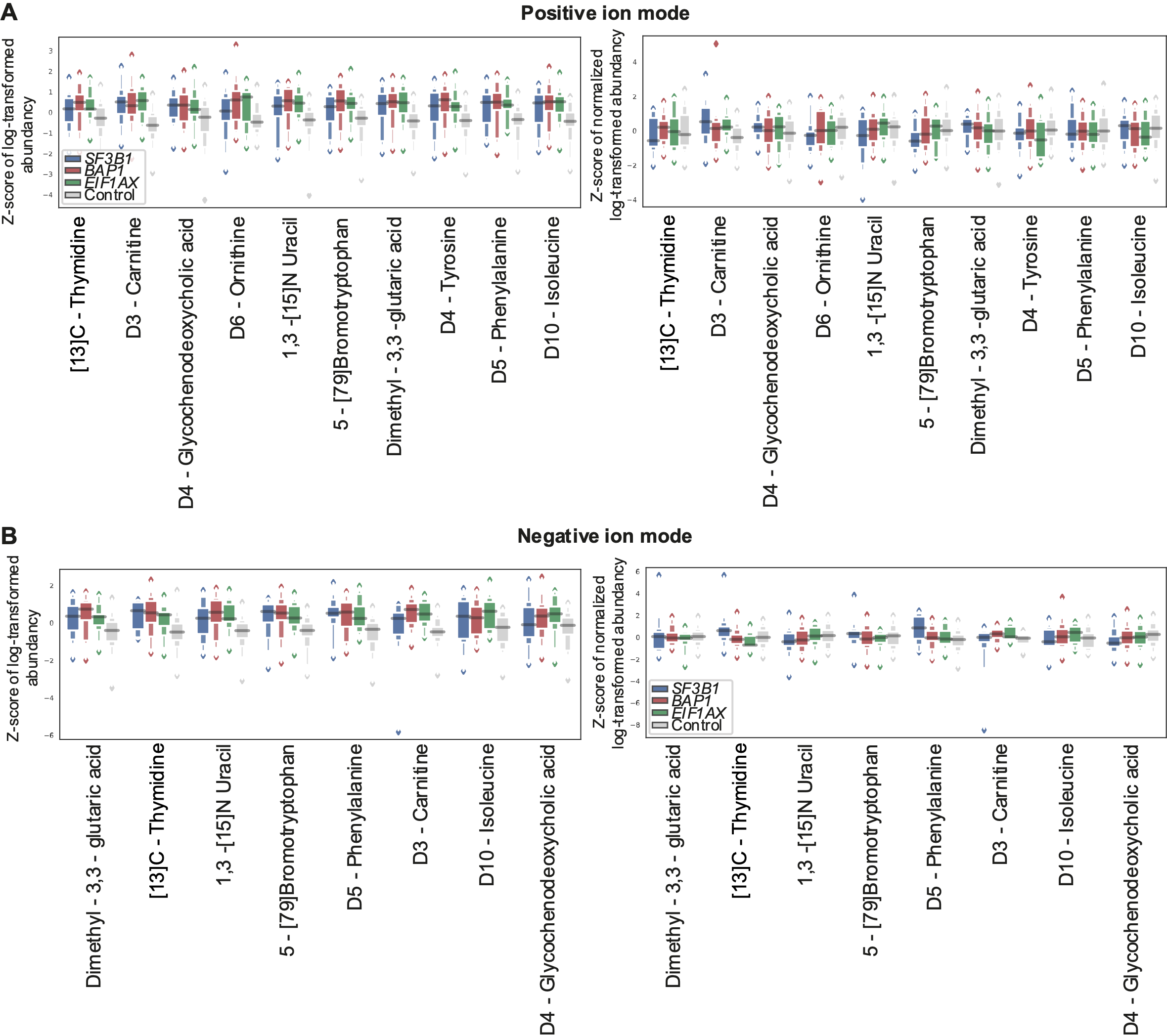

**eFigure 2. Quality control characteristics by internal standards in discovery set.**

To eliminate technical variances, internal standards were added to each sample. These internal standards were used to normalize feature abundancies (see eMethods). For some groups we observe overall abundancy differences for the internal standards. Normalization reduced these group differences when looking at the normalized internal standards. Boxplots show z-scores of log-transformed abundancies of the internal standards in the discovery set in the positive ion mode (A) and negative ion mode (B); on the left are unnormalized abundancies and the right side shows normalized abundancies.

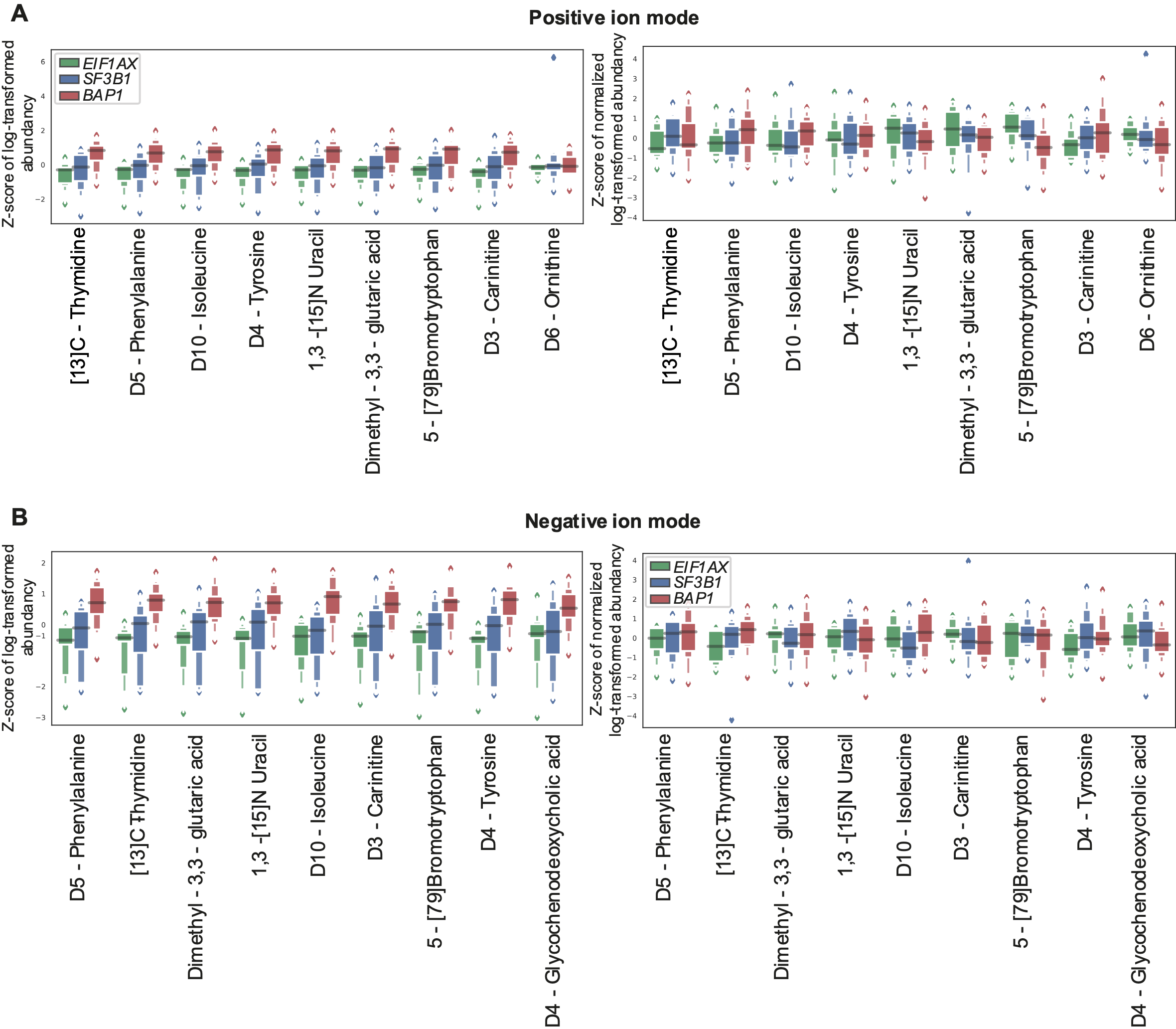
**eFigure 3. Quality control characteristics by internals standards in replication set.**

To eliminate technical variances in the replication set, internal standards were used to normalize feature abundancies. After normalization, technical differences in metabolite abundancy were reduced in the (A) positive ion mode and (B) negative ion mode.

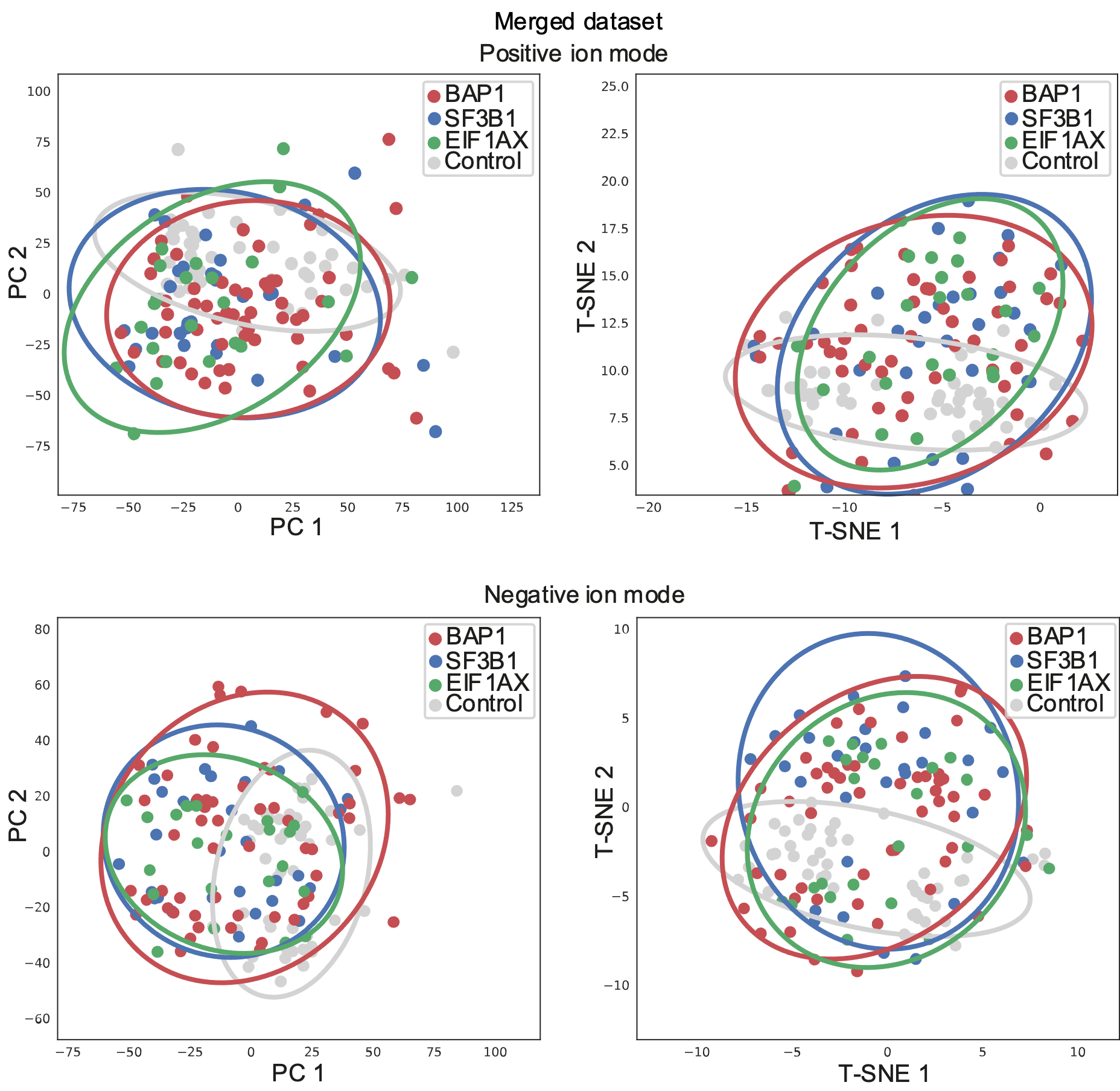

**eFigure 4.** **Quality control by two-dimensionality reduction analyses.** Unsupervised clustering by principal component analysis (PCA) and T-distributed Stochastic Neighbor Embedding (T-SNE) on all normalized feature abundancies show no deviations in (normalized) feature abundancies between plasma samples of patients with *BAP1*, *SF3B1* or *EIF1AX* and control-participants in the whole cohort consisting of the discovery and replication sets (i.e., the merged dataset) in positive and negative ion modes.

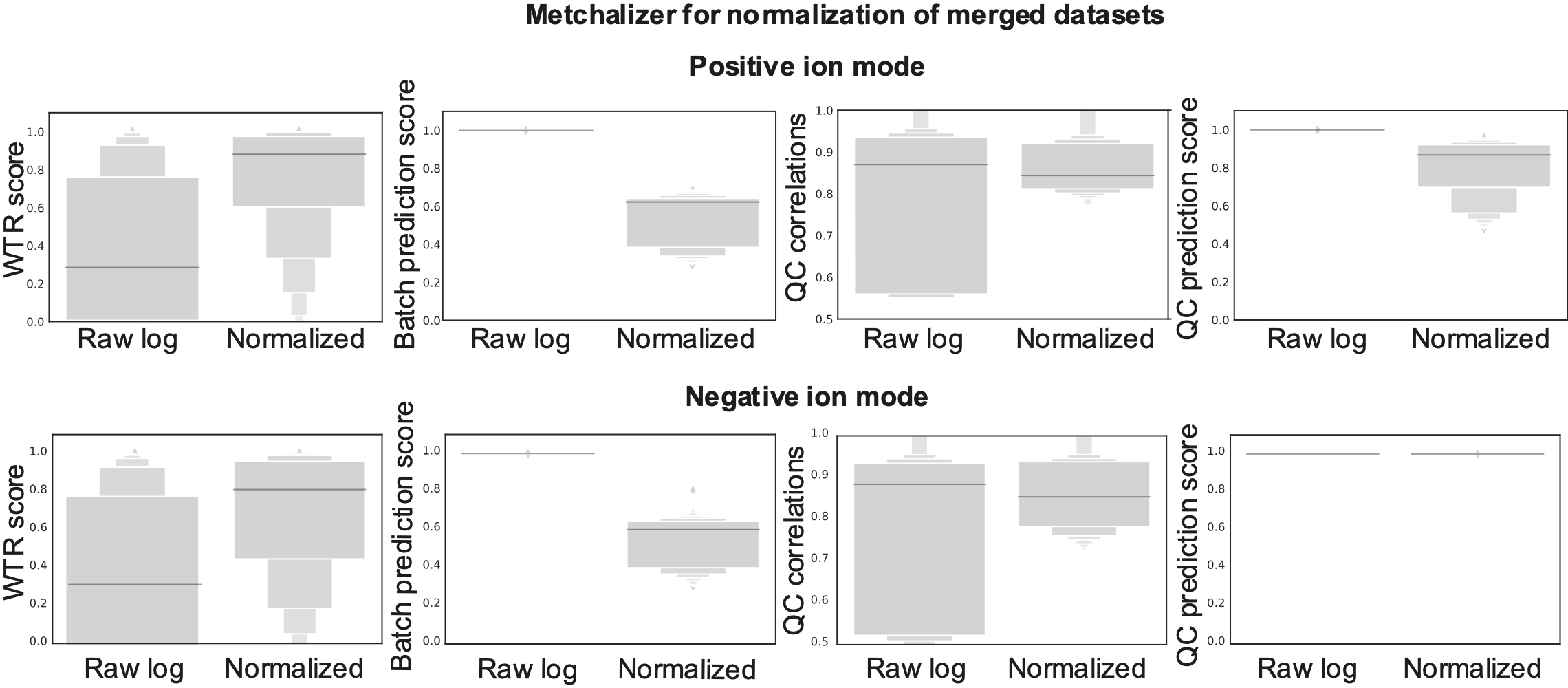
**eFigure 5. Assessment of Metchalizer normalization in the merged dataset.**

The four metrics WTR scores, Batch prediction scores, QC correlations and QC prediction scores are determined for both the log-transformed raw data and the Metchalizer normalized data. Technical replicates of the same sample were included in the replication and discovery set. Additionally, technical replicates were included in each individual dataset. Assessment of these technical replicates indicate improved normalization as they are associated with increased WTR scores, QC correlations and QC predictions scores, and with reduced batch prediction scores^16^. These metrics are determined for both ion modes.

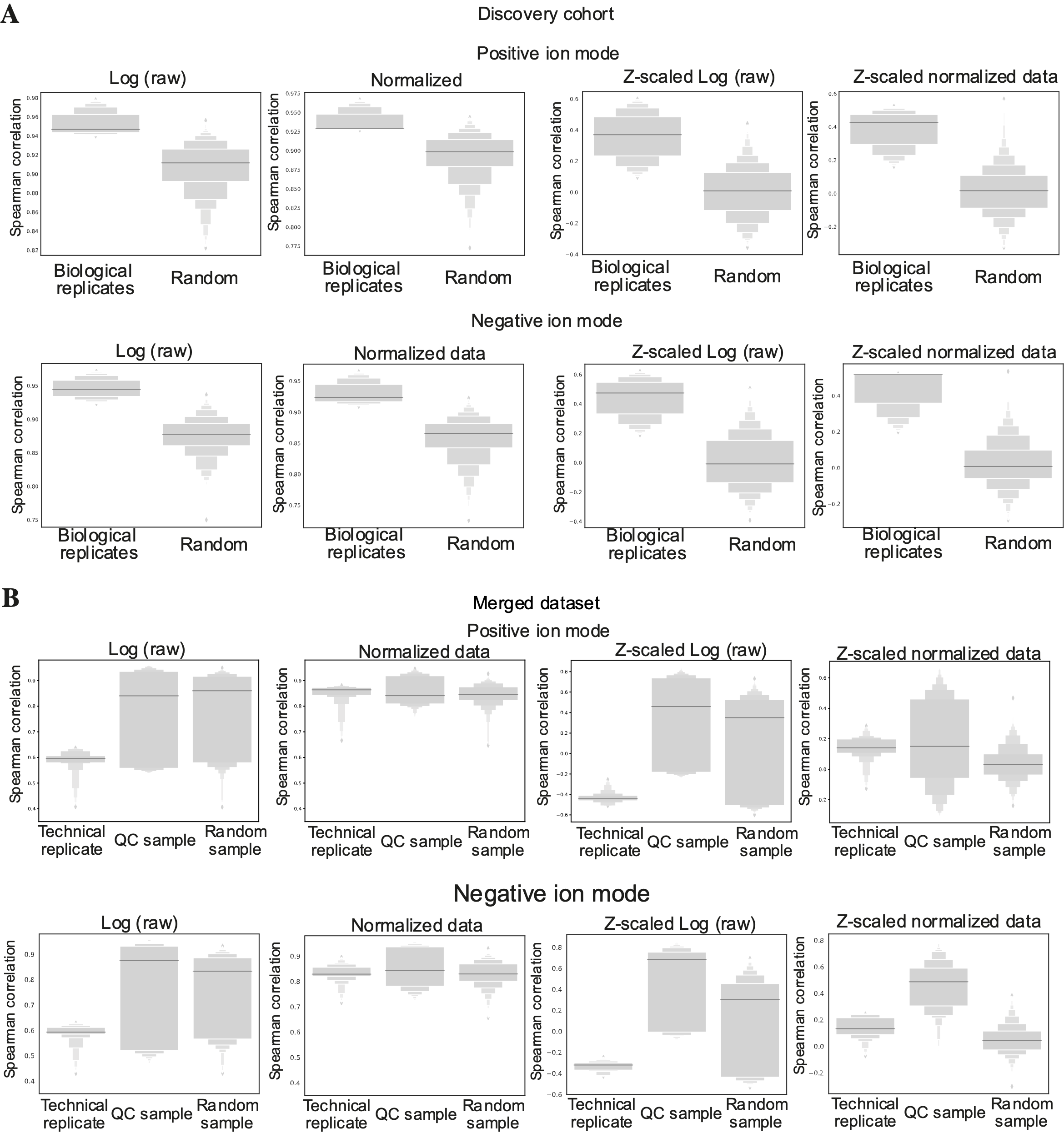
**eFigure 6. Assessment of technical and biological variation by using replicates in the discovery cohort and merged dataset.**

The Spearman correlation between two samples was determined using all features for samples in the discovery cohort, where abundancies were either log-transformed raw abundancies or normalized abundancies. These correlations were determined between biological replicates (two different UM-patient plasma samples retrieved at one time-point), or between a random selection of samples. For the latter we expect, and observe, lower correlation coefficients than the biological replicates. Furthermore, normalized abundancies should ideally further improve the correlation between the latter (A). The Spearman correlation between two samples was determined using all features for samples in the merged dataset, where abundancies were either log-transformed raw abundancies or normalized abundancies. Correlations were determined between technical replicates (a UM-patient sample being measured in each of the two batches), between the QC samples (same sample measured multiple times per batch and in both batches) and between a random selection of samples (B). The Spearman correlation of normalized feature abundancies of biological and technical replicates (both UM-patient samples and QC-samples) showed better correlation than randomly chosen samples.

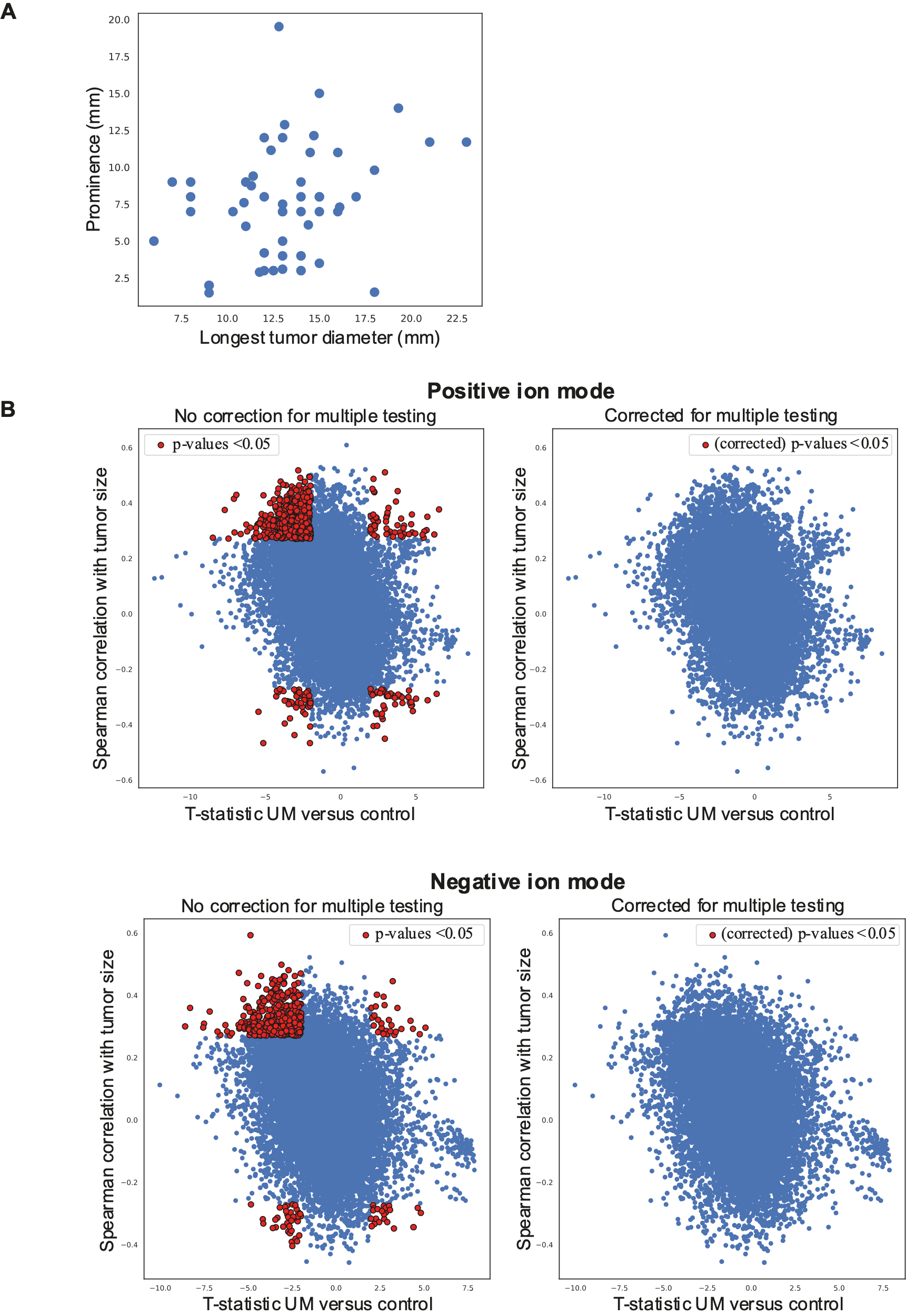

**eFigure 7. Correlation of m/z features with tumor size and differential abundance between UM patients and controls.**

Since tumor size characteristics prominence and longest tumor diameter (LTD) correlate (A), the first PC from PCA analysis was used as metric to correlate with feature abundance. Next, the t-statistic of feature abundance in UM-patients versus control-participants was calculated. By scattering both metrics, we obtain interesting regions in each corner of the scatter plot (B) and these features are depicted by red dots (p-value <0.05). In the top-right corner of the plot the relatively high feature abundance in UM-patients and (strong) correlation with tumor size are depicted; the bottom-left corner shows low feature abundance in UM-patients and inverse correlation to tumor size. No feature is significantly differentially abundant between UM-patients and control-participants and correlated with tumor size, after correcting for multiple testing using the Benjamini/Hochberg procedure with a family-wise error rate of 0.05.

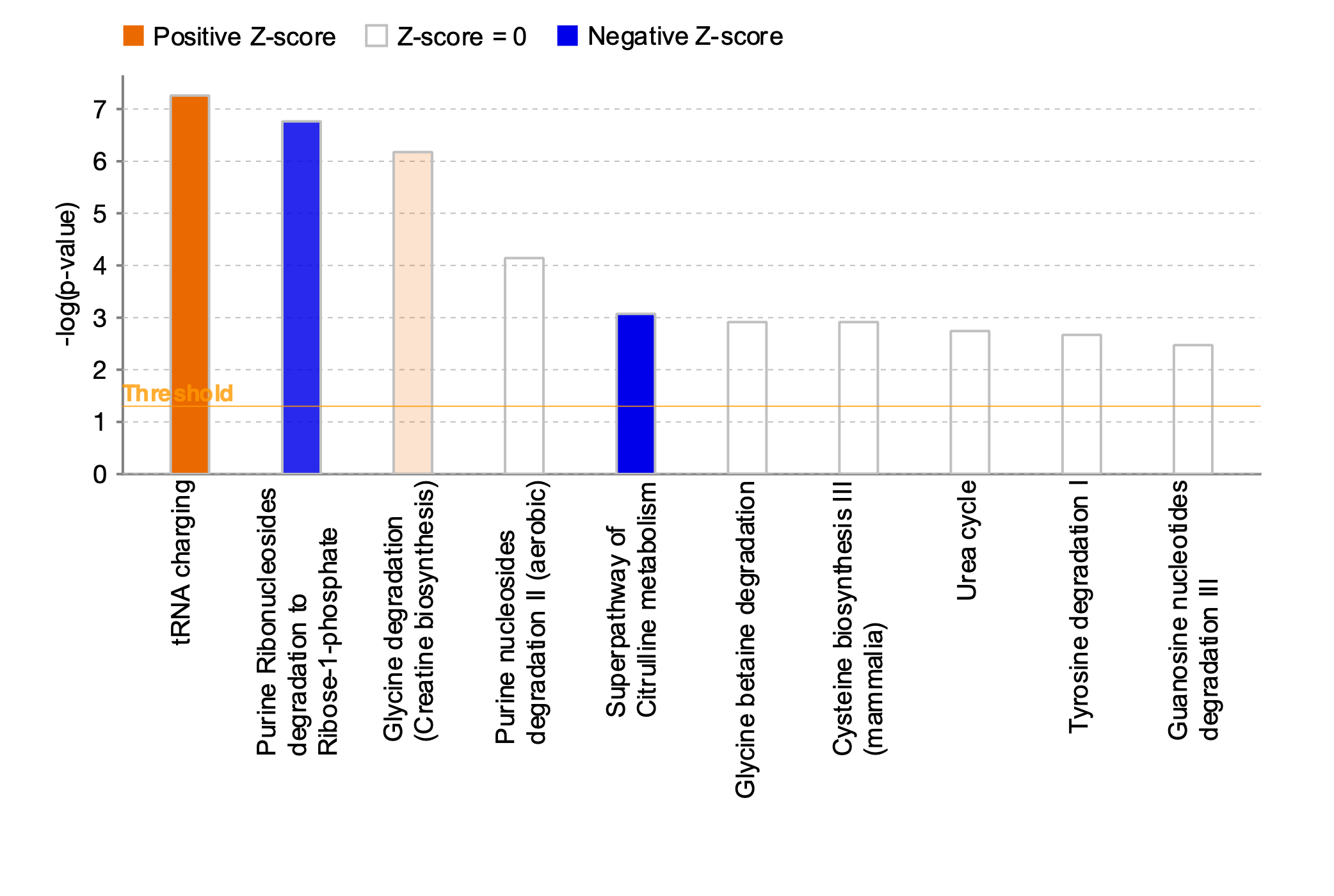

**eFigure 8.** **Pathway analysis of top-10 affected pathways in UM-patients compared to control-participants.**

All metabolite annotations with a HMDB ID were pulled from both ion modes. For metabolites that were annotated in both ion modes, we selected the ion mode that had on average the highest abundancy (i.e., better signal-to-noise ratio). The fold change between UM-patients and control-participants and the p-values from the t-test were used as input for Ingenuity Pathway Analysis (IPA). Metabolites were annotated using an in-house developed database as described previously^17^. Data was analyzed using QIAGEN IPA (QIAGEN Inc., <https://digitalinsights.qiagen.com/IPA>). Z-scores were obtained by calculating the Fold Change between annotated metabolite abundancy in UM-patients compared to control-participants. The p-values were obtained by t-test of differences in metabolite abundancy between UM-patients and control-participants. Upregulation of pathways is depicted in orange and downregulation is visualized by blue bars, in which color intensity correlates to the Z-score. We observed an upregulation of transfer (tRNA) charging and the use of glycine for the creatine biosynthesis. Purine ribonucleosides degradation and the super pathway of citrulline metabolism were downregulated in UM-patients. tRNA charging generally promotes oncogenesis^18^ and, consistent with upregulation of tRNA charging, we observed a lower abundancy of arginine, asparagine, cysteine, leucine, lysine, methionine, phenylalanine, proline, tryptophan, tyrosine and valine in UM-patients. Extracellular glycine is internalized by cells and degraded to produce substrates for nucleotides, proteins, gluthatione and methylation^19^*.* Additionally, we observed a lower abundance of adenine, guanine, and D-ribose-5-phosphate, which indicate a downregulation of purine degradation or increased salvage of bases and can indicate a higher energy consumption. Guanine is metabolized for recycling bases by hypoxanthine-guanine phosphoribosyltransferase that metabolizes guanine into guanine-monophosphate (GMP) and the purine xanthine. An upregulation of this purine salvage aids cell proliferation^20^. Arginine is lower abundant that can be metabolized for the production of nitric oxide and citrulline^21^. Nitric oxide that is obtained from arginine by the citrulline metabolism can regulate cell death, angiogenesis and proliferation^22^.

**Supplementary tables**

| **Primary driver** | **Secondary driver mutation** | **Copy number variations** | **Metastatic risk** | **Typical metastatic-free survival** |
| --- | --- | --- | --- | --- |
| G-protein coupled receptor pathway:  *GNA11/GNAQ/*  *CYSLTR2/PLCB4* | *BAP1* | Large, whole chromosomes or arms: chr. 3 loss; 1p loss; 8 ,8q gain | High | < 5 years |
|  | *SF3B1* | Small, structural changes: 1p loss; 11q loss; 6p gain; 8q gain | Intermediate | bimodal: <5 or > 7 years |
|  | *EIF1AX* | chr. 6 gain; 6p gain | Low | no metastases |

**eTable 1.** **Secondary driver mutation responsible for malignant transformation and large prognostic factor, together with copy number aberrations.** Loss-of-function mutations in tumor suppressor gene BRCA associated protein 1 (*BAP1*) are correlated with large structural chromosomal aberrations, primarily loss of chromosome 3 and 1p and 6p or 8q isochromosome formation with concurrent loss of chromosome 6q and gain of chromosome 8q. These tumors are associated with a metastatic free survival of less than five years^23^. Splicing factor 3 subunit B1 (*SF3B1*) is involved in the spliceosome complex, *SF3B1*-mutated UMs frequently have smaller telomeric chromosomal aberrations and partial 8q gain. These tumors pose an intermediate risk of metastasizing and metastases arise within 5 to 15 years^24^. Eukaryotic translation initiation factor 1A, X-linked (*EIF1AX*)-mutated tumors usually occur with a concurrent gain of chromosome 6p and these tumors hardly ever metastasize^24^. The *SF3B1* and *EIF1AX* secondary driver mutations occur with disomy 3 in UM^23,24^

| **Study ID** | **Sex** | **Age at phlebotomy (range in years)** | **Disease** |
| --- | --- | --- | --- |
| **11603** | Male | 76-80 | AMD |
| **11621** | Male | 76-80 | AMD |
| **11628** | Male | 71-75 | Cataract |
| **11643** | Female | 61-65 | AMD |
| **11652** | Female | 76-80 | AMD |
| **11657** | Female | 61-65 | Cataract |
| **11687** | Female | 86-90 | Branch retinal vein occlusion |
| **11743** | Female | 81-85 | AMD |
| **11794** | Male | 81-85 | Traumatic cataract |
| **11819** | Female | 56-60 | Open angle glaucoma |
| **11825** | Male | 61-65 | AMD |
| **11826** | Male | 76-80 | AMD |
| **11828** | Female | 86-90 | AMD |
| **11900** | Female | 61-65 | Cataract |
| **11901** | Female | 61-65 | Cataract |
| **11932** | Male | 81-85 | AMD |
| **11952** | Male | 71-75 | Cataract |
| **11963** | Female | 65-70 | Cataract |
| **11965** | Male | 81-85 | Cataract |
| **11966** | Male | 81-85 | Cataract |
| **11967** | Male | 96-100 | AMD |
| **11969** | Male | 81-85 | Diabetic macular edema |
| **11970** | Female | 56-60 | Cataract |
| **11971** | Male | 66-70 | Cataract |
| **11972** | Male | 81-85 | Cataract |
| **11978** | Female | 76-80 | AMD |
| **12156** | Male | 65-70 | AMD |
| **12157** | Male | 65-70 | Cataract |
| **12158** | Female | 71-75 | Cataract |
| **12162** | Female | 61-65 | Cataract |
| **12163** | Female | 66-70 | Cataract |
| **12165** | Female | 81-85 | AMD |
| **12166** | Male | 71-75 | Cataract |
| **12167** | Male | 61-65 | Cataract |
| **12168** | Female | 61-65 | AMD |
| **12175** | Female | 86-90 | AMD |
| **12178** | Female | 71-75 | AMD |
| **12237** | Male | 76-80 | Cataract |
| **12280** | Female | 61-65 | Cataract |
| **12287** | Female | 86-90 | AMD |
| **12288** | Female | 80-85 | AMD |
| **12291** | Female | 76-80 | AMD |
| **12292** | Male | 76-80 | Cataract |
| **12293** | Female | 71-75 | Central retinal vein occlusion |
| **12294** | Male | 71-75 | AMD |
| **12296** | Female | 66-70 | Cataract |

**eTable 2. Characteristics of control-participants.**

Control-participants are included from the Combined Ophthalmic Research Rotterdam Biobank (CORRBI). These patients had blood drawn prior to surgical treatment and no systemic treatment was administered during the period of phlebotomy. Abbreviation: AMD: Age-related macular degeneration. Shown study identification numbers are only traceable by the principle investigator and specifically authorized members of the research group.

| **Sex** | **Age at onset (range in years)** | **DFS (months)** | **Plasma retrieval** | **Mutation** | **Replication set** |
| --- | --- | --- | --- | --- | --- |
| Male | 56-60 | 95,7 | 1998 | *SF3B1* | Technical replication |
| Male | 66-70 | 8,2 | 2003 | *BAP1* | Technical replication |
| Male | 61-65 | 23,7 | 2004 | *BAP1* | Technical replication |
| Female | 71-75 | 19,4 | 2004 | *EIF1AX* | Technical replication |
| Female | 71-75 | 19,4 | 2004 | *EIF1AX* | Biological replication |
| Female | 26-30 | 154,1 | 2006 | *SF3B1* | Technical replication |
| Female | 71-75 | 130,9 | 2008 | *SF3B1* | Technical replication |
| Female | 66-70 | 143,8 | 2008 | *EIF1AX* | Technical replication |
| Male | 81-85 | 63,2 | 2008 | *EIF1AX* | Technical replication |
| Female | 56-60 | 15,8 | 2009 | *BAP1* | Technical replication |
| Male | 66-70 | 48,5 | 2010 | *BAP1* | Technical replication |
| Female | 71-75 | 102,3 | 2011 | *EIF1AX* | Technical replication |
| Female | 41-45 | 67,5 | 2014 | *SF3B1* | Technical replication |
| Male | 46-50 | 70 | 2015 | *SF3B1* | Technical replication |
| Male | 46-50 | 70 | 2015 | *SF3B1* | Biological replication |
| Female | 46-50 | 11,3 | 2018 | *SF3B1* | Technical replication |
| Female | 46-50 | 11,3 | 2018 | *SF3B1* | Biological replication |
| Male | 55-60 | 14,3 | 2019 | *EIF1AX* | Technical replication |

**eTable 3. Patient and sample characteristics of the technical and biological replication sets.**

To compare metabolite profiles and evaluate batch-effects a technical replication set comprising of 15 samples was analyzed in both the discovery and replication set. Additionally, three samples were used as a biological replicate in which a different plasma sample was retrieved at the same time-point. The patients resemble their respective cohorts in metastatic free survival (MFS), age at onset and male/female ratio. These samples were retrieved and subsequently stored between 1998 and 2019.

| A | **Discovery cohort UM-subtypes and control-participants** | | | | | |  |
| --- | --- | --- | --- | --- | --- | --- | --- |
|  | **Negative ion mode** | | |  | **Positive ion mode** | | |
|  | precision | recall | F1-score |  | precision | recall | F1-score |
| *BAP1* | 0.532 | 0.676 | 0.595 | *BAP1* | 0.453 | 0.649 | 0.533 |
| *SF3B1* | 0.000 | 0.000 | 0.000 | *SF3B1* | 0.000 | 0.000 | 0.000 |
| *EIF1AX* | 0.000 | 0.000 | 0.000 | *EIF1AX* | 0.000 | 0.000 | 0.000 |
| Control | 0.714 | 0.978 | 0.826 | Control | 0.800 | 0.957 | 0.871 |
| Accuracy | 0.614 |  |  | Accuracy | 0.600 |  |  |
| B | **Replication cohort UM-subtypes** | | | | | |  |
|  | **Negative ion mode** | | |  | **Positive ion mode** | | |
|  | precision | recall | F1-score |  | precision | recall | F1-score |
| *BAP1* | 0.652 | 0.790 | 0.714 | *BAP1* | 0.700 | 0.778 | 0.737 |
| *SF3B1* | 0.556 | 0.625 | 0.588 | *SF3B1* | 0.474 | 0.563 | 0.514 |
| *EIF1AX* | 0.000 | 0.000 | 0.000 | *EIF1AX* | 0.750 | 0.333 | 0.462 |
| Accuracy | 0.568 |  |  | Accuracy | 0.605 |  |  |
| C | **Merged datasets UM-subtypes and control-participants** | | | | | |  |
|  | **Negative ion mode** | | |  | **Positive ion mode** | | |
|  | precision | recall | F1-score |  | precision | recall | F1-score |
| *BAP1* | 0.565 | 0.64 | 0.600 | *BAP1* | 0.581 | 0.705 | 0.637 |
| *SF3B1* | 0.367 | 0.289 | 0.324 | *SF3B1* | 0.500 | 0.289 | 0.367 |
| *EIF1AX* | 0.410 | 0.321 | 0.360 | *EIF1AX* | 0.444 | 0.286 | 0.348 |
| Control | 0.780 | 1.000 | 0.876 | Control | 0.763 | 0.978 | 0.857 |
| Accuracy | 0.601 |  |  | Accuracy | 0.618 |  |  |

**eTable 4.** **No differentially abundant metabolites in plasma of UM-patients between subtypes.**

Leave-one-out cross-validation of a Random Forest classifier (RFC) trained on all features for distinguishing samples based on secondary driver mutation and mutation status. F1-scores resemble the harmonic mean of precision and recall and are thus the metric for combined performance of the classifier. The trained RFC performs poorly in the discovery cohort, meaning that a distinction of secondary driver mutation cannot be made based on m/z features with F1-scores of 0.60, 0.00 and 0.00 for patients harboring a *BAP1*, *SF3B1* and *EIF1AX*-mutated tumor in the negative ion mode, respectively; and 0.53, 0.00 and 0.00 for patients harboring a *BAP1*, *SF3B1* and *EIF1AX*-mutated tumor in the positive ion mode, respectively (A). In the replication cohort, abundancies of m/z features are slightly different in the *BAP1* group (B) with F1-scores of 0.82, 0.59 and 0.59 for patients harboring a *BAP1*, *SF3B1* and *EIF1AX*-mutated tumor in the negative ion mode, respectively. In the positive ion mode F1-scores were 0.65, 0.35 and 0.64 for patients harboring a *BAP1*, *SF3B1* and *EIF1AX*-mutated tumor, respectively. These differences did not hold true in the meta-analysis using the merged dataset (C). However, controls are detected, even when the three mutational groups are selected showing differential feature abundancies.

| A | **Merged datasets primary driver mutation** | | | | | |  |
| --- | --- | --- | --- | --- | --- | --- | --- |
|  | **Negative ion mode** | | |  | **Positive ion mode** | | |
|  | precision | recall | F1-score |  | precision | recall | F1-score |
| *GNA11* | 0.571 | 0.571 | 0.571 | *GNA11* | 0.571 | 0.571 | 0.571 |
| *GNAQ* | 0.609 | 0.609 | 0.609 | *GNAQ* | 0.609 | 0.609 | 0.609 |
| Accuracy | 0.591 |  |  | Accuracy | 0.591 |  |  |
| B | **Merged dataset *BAP1* and *EIF1AX* mutations** | | | | | |  |
|  | **Negative ion mode** | | |  | **Positive ion mode** | | |
|  | precision | recall | F1-score |  | precision | recall | F1-score |
| *BAP1* | 0.571 | 0.632 | 0.600 | *BAP1* | 0.636 | 0.778 | 0.700 |
| *EIF1AX* | 0.417 | 0.357 | 0.385 | *EIF1AX* | 0.000 | 0.000 | 0.000 |
| Accuracy | 0.515 |  |  | Accuracy | 0.538 |  |  |
| C | **Merged dataset metastatic formation of UM-patients** | | | | | |  |
|  | **Negative ion mode** | | |  | **Positive ion mode** | | |
|  | precision | recall | F1-score |  | precision | recall | F1-score |
| Yes | 0.500 | 0.150 | 0.231 | Yes | 0.333 | 0.100 | 0.154 |
| No | 0.726 | 0.938 | 0.818 | No | 0.710 | 0.917 | 0.800 |
| Accuracy | 0.706 |  |  | Accuracy | 0.676 |  |  |

**eTable 5. No differences in metabolite profiles associated with poor prognosis.**

In the primary driver gene *GNA11,* mutations are associated with a shorter metastatic free survival and a loss of BAP1 expression^25^. We deployed a Random Forest classifier using a leave-one-out approach in the merged dataset and could not detect differential feature patterns between *GNAQ* and *GNA11* (A). Furthermore, converse to the supervised discriminant analysis (eFigure 6), this more robust, leave-one-out cross-validation approach does not show differences in metabolite patterns between patients harboring a *BAP1* or *EIF1AX*-tumor (i.e., tumors with poor and good prognosis) using the merged dataset (B). Ultimately, an RFC was trained to classify patients based on the future development of metastases. We did not detect differences in metabolite patterns between patients who would prove to develop metastases and patients without metastatic formation after at least 60 months of follow-up (the timepoint at which most metastases have presented). The leave-one-out cross-validation of the trained RFC shows decent performance when classifying non-metastatic patients, however premetastatic patients could not be classified based on metabolite profiles (C).

**eMethods**

### **Patient selection**

Patients were selected based on their secondary driver mutation (*EIF1AX* (n=24), *SF3B1* (n=33) or *BAP1* (n=53)) and availability of peripheral blood prior to treatment and they were age and sex-matched. Survival data and tumor characteristics are recorded; additionally, liver function tests and medical imaging is performed routinely at intervals of six months for the screening of (hepatic) metastases. Furthermore, known confounding diseases (such as cardiovascular disease, diabetes or pulmonary disease) were collected and noted from patient information next to medication use (data not shown).

The patients enrolled in the Rotterdam Ocular Melanoma Study group (ROMS) between 1998 and 2021 (METC: MEC-2009-375) (Table 1). Treatment for the primary tumor and phlebotomy were performed at Erasmus MC (Rotterdam, the Netherlands) or the Rotterdam Eye Hospital (Rotterdam, the Netherlands). Age and sex-matched non-UM controls (n=46) consisted of patients receiving care for ophthalmic diseases (eTable 2) in Erasmus MC and had peripheral blood withdrawn prior to treatment between 2016 and 2017 (Biobank Combined Ophthalmic Research Rotterdam Biobank (CORRBI) (METC: MEC-2012-031)) (Table 1).

### **Collection of blood**

Blood was collected in 8ml Lithium-Heparin blood collection tubes between 1998 and 2021. Within 6 hours after blood collection, plasma was separated from red and white blood cells by centrifuging once at 3500g at 4°C for 10 minutes. The supernatant was centrifuged a second time at 17000g at 4°C for 10 minutes and subsequently the supernatant was collected and stored at -80 °C. The average storage time per sample did not differ between the individual groups of mutations and controls.

### **Liquid chromatography-mass spectrometry**

Ultra-high performance liquid chromatography (UHPLC) Orbitrap mass-spectrometry was performed on plasma of peripheral blood, as described in R. Bonte *et al.*^17^. UHPLC-MS analyses were performed using a Dionex Ultimate 3000 UHPLC chromatograph and Q Exactive Plus hybrid quadrupole-Orbitrap mass spectrometer with heated electrospray source. The injection volume was 3 µL for all samples. A flow rate of 400 µL/min was used for chromatographic elution. UHPLC-MS sample analyses were performed in positive and negative ion modes as some metabolites are only detected in one of the two modes. Capillary voltage in the negative ion mode was set to -3.5 kV and 3.5 kV in the positive ion mode and the capillary temperature was set at 380 ˚C and auxiliary temperature was set at 300 ˚C during analyses. UHPLC sample analyses in both positive and negation ion modes were preceded by four injections of water and a quality control sample; hereafter the samples were run in a randomized order.

To eliminate systemic variation, internal and external standards mixtures were added, and feature abundance was normalized on the abundance of these internal and external standards. The internal mixture contains: D5 - L-Phenylalanine (600 µmol/L), [^13^C] - Thymidine (300 µmol/L), 1,3-^15^N -Uracil (300 µmol/L), D10 - Isoleucine (500 µmol/L), D6 - Ornitihine (225 µmol/L), D4 - Tyrosine (230 µmol/L), 5-bromo-DL-tryptophan (85 µmol/L), 3,3-dimethylglutaric acid (300 µmol/L), D4 - glycochenodeoxycholic acid (44 µmol/L), D3 - Carnitine (285 µmol/L). The external mixture contains D3 - methylmalonic acid (100 µmol/L), D2 - Uridine (207 µmol/L), D8 - L-valine (670 µmol/L), D2 - acetylcarnitine (45 µmol/L), D3 - hexanoylcarnitine (19 µmol/L), D3 - tetradecanoylcarnitine (19 µmol/L), D3 - hexadecanoylcarnitine (6 µmol/L).

### **Metabolomics analyses**

All feature abundancies were log-transformed to log(1+x) and Z-transformed (after normalization) after which they followed a Gaussian distribution. All non-zero values were pooled (across all features and samples). From this pool the 2^nd^ percentile was determined. A feature was included when at least 5 samples had an abundancy above the 2^nd^ percentile. Furthermore, for the merged dataset only features were considered that were detected in both batches.

**Removing inter - and intra-batch variation**

Two different normalization methods were used to correct for inter- and intra-batch variations. For the analysis of the replication - and discovery set, all features were normalized using a linear regression (BayesianRidge method from Scikit-learn^26^) on the first two principal components of the internal standards. This results in abundancies that are corrected by the overall abundancies of the internal standards. However, this normalization step was only performed on features where 75% of the samples had an abundancy above zero. Furthermore, prior to fitting the model outlier samples were removed when sample |Z-score| > 3. Note that features for which less than 75% of the samples had an abundancy above zero were not normalized, and thus their original (log-transformed) abundancy was used in the analysis.

For the merged dataset, we applied Metchalizer (v1, https://github.com/mbongaerts/Metchalizer) with an initial log-transformation (log(x+1)). Four metrics were used to judge the performance of normalization: WTR-score (Within variance Total variance Ratio), quality control correlations, batch prediction score and QC prediction score, as described by Bongaerts *et al.*^16^.

#### **Statistical analyses**

For unsupervised clustering, the PCA and T-SNE dimensionality reduction methods from Scikit-learn were used to explore potential clustering of the four groups. Afterwards for supervised clustering the Partial Least-Squares Determinant Analysis from Scikit-learn (PLS-DA) was used to assess clustering of the four groups based on m/z feature abundance. A Random Forest Classifier (RFC) from Scikit-learn^26^ containing 150 decision trees and a maximum depth of 100 was trained for classifying UM-patients versus non-UM controls and for classifying the individual secondary mutation groups based on m/z feature abundancy. We used a leave-one-out cross-validation strategy to assess the performance of the classifier. For each validation round, one sample from the dataset was left out and put into the test set. Since not each group is equally represented in the dataset, classification biases might occur for groups having more samples. To correct for this potential bias, we oversampled each group such that each group had an equal number of samples (n_i_=200). Normal noise N(0, 0.25) was added to the dataset to prevent overfitting, that serves as regularization. For each sample we obtained a ‘probability score’ of the correct class using the predict_proba() method from the RFC. ROC curves for UM-patients versus controls-participants were created using these ‘probability scores’. In order to estimate the variance on the area under the curve (AUC) a bootstrap with replacement was used. In total 25 bootstraps were performed, from which the average – and standard deviation on the AUC was determined. Ingenuity Pathway Analysis (version 01-20-04) was used for analyzing differentially regulated pathways using annotated metabolites^27^.
